# How do scientific evidence and clinical experience influence physiotherapists’ confidence in a treatment’s efficacy ? A survey of French-speaking physiotherapists

**DOI:** 10.1101/2025.07.26.25331629

**Authors:** Nicolas Pinsault, François Générau

## Abstract

**Background and Objectives:** Current physiotherapy practice is not always in line with the best available scientific evidence. Some barriers to evidence-based practice have been identified with self-declared questionnaires, but it remains unknown how much physiotherapists agree that high-quality scientific evidence is more reliable than clinical experience to evaluate a treatment’s efficacy. The goal of this study is to investigate the relative influence of research findings, positive and negative, and clinical experience on physiotherapists’ confidence in a treatment’s efficacy.

**Materials and Method:** In a web-based survey, French-speaking physiotherapists in France and Belgium and physiotherapy students gave their degree of confidence in the efficacy of four fictitious treatments whose characteristics varied regarding scientific evidence and clinical experience, on a 0-100 scale.

**Results:** We found that 20.9 % of students (95%CI [16.4, 26.2]) and 17.3 % of physiotherapists (95%CI [13.1, 22.4]) had equal or higher confidence in clinical experience than scientific evidence. Moreover, for about a fourth of the participants, negative placebo-controlled trial findings had no impact on their confidence in a treatment’s efficacy, while for about half of the participants, such findings *increased* their confidence in a treatment’s efficacy.

Level of training in evidence-based practice seems to have a null to at most moderate effect on the relative confidence in scientific evidence and clinical experience.

**Conclusion:** High confidence in clinical experience may have been an underestimated barrier to evidence-based practice in previous studies. The interpretation of negative findings of placebo-controlled trials show that most physiotherapists misunderstand placebos or ignore negative study findings. This could also be an important barrier to evidence-based practice.

## Introduction

### Low-value care in physiotherapy

Following Scott and Duckett (1), low-value care may be defined as an intervention where evidence suggests it confers no or very little benefit on patients, or risk of harm exceeds likely benefit, or, more broadly, the added costs of the intervention do not provide proportional added benefits.

Evidence indicates that low-value care is prevalent in all healthcare systems worldwide (2,3), and physiotherapy is no exception. For instance, in a recent systematic review (4), the authors found that “across all musculoskeletal conditions, 54% of physical therapists chose recommended treatments, 43% chose treatments that were not recommended and 81% chose treatments that have no recommendation (based on surveys completed by physical therapists)”. What is more, the proportion of physiotherapists providing not recommended treatments didn’t seem to decrease between 1990 and 2018 (5).

### Barriers to evidence-based practice

So why isn’t current physiotherapy practice always based on the best evidence ? The concept of evidence-based practice (EBP) has been recognized as fundamental in healthcare in general for at least twenty years (6). In France, the physiotherapists’ code of ethics specifies that care should be based on scientific data since 2008 (7).

Some effort has been made to identify the barriers to EBP, in healthcare and in physiotherapy in particular (8– 11). In the systematic review (8) on physiotherapy, identified barriers were “related to the organizational context (lack of time, access and support), education (language, lack of research and statistical skills), personal behavior (lack of interest) and limits of EBP (lack of generalizability)”. However, the lack of interest was a rarely declared barrier. Moreover, studies that investigated the attitudes of physiotherapists towards EBP have generally reported positive average attitudes (12). In France, in particular, a recent study found that EBP was perceived by its sample of physiotherapists as relevant to their practice (average score of 80/100 in the relevance domain of the EBP^2^ questionnaire) (13). However, it should be noted that these studies are based on self-declared questionnaires, so we may expect a social desirability bias in the results, in favor of EBP. All in all, to the best of our knowledge, it is unknown how much physiotherapists actually consider that scientific evidence is a much more robust way of evaluating a treatment’s efficacy than clinical experience.

Our goal is to investigate the relative importance of clinical experience and scientific evidence, positive or negative, in physiotherapists’ confidence in treatments’ efficacy. In the following, ‘negative evidence’ should be understood as evidence for the absence of efficacy of a treatment.

## Materials and Methods

We conducted a cross-sectional web-based survey, among a convenience sample of French-speaking physiotherapists and physiotherapy students.

### Survey description

We built the survey into two parts: a main question and some demographic questions. The translated English version of the questionnaire is provided in the supplementary material (SM1) and the original French version is available on request. The survey was pretested on a sample of 5 physiotherapists and 5 physiotherapy students from different years of study, to check for adequate understanding of all the questions. This phase showed that the cognitive load of the main question was high. It allowed us to simplify the question as much as possible, and include a warning so that the participants would read and answer the question carefully.

#### Main question

The complete English translation of the main question is given in the Text Box below. A written brief generic clinical situation is described to the participant, where they may use one of four treatment options. Each of the four treatments has particular characteristics regarding scientific evidence and clinical experience, which are summarized in Table 1.

**Table 1:** Characteristics of the four treatment options of the main question, regarding clinical experience and scientific literature.

|  | Clinical experience | Scientific literature |
| --- | --- | --- |
| Treatment A | Positive clinical experience | Non-existent |
| Treatment B | Positive clinical experience | Negative evidence |
| Treatment C | None | Positive evidence |
| Treatment D | None | Non-existent |

Text box: English translation of the main question

You are treating a new patient, Mr. X. To treat his health problem, you are considering 4 possible treatments (A, B, C and D), which have the following characteristics.

**Treatments A and B**: These have long been used in your profession. You are accustomed to using one or the other, and you observe an improvement in the condition of the patients with whom you use these treatments. What’s more, your colleagues also use them and are satisfied. However, one of them recently pointed out to you that there are no studies to prove their effectiveness. On further enquiry, you discovered the following facts:

- Regarding **treatment A**, there are several very high-quality, independent studies comparing it to placebo. These have not demonstrated superior efficacy to placebo.
- Regarding **treatment B**, there are no studies measuring its efficacy.

**Treatments C and D**: A colleague recently told you about these two options, which, according to some sources, are showing interesting results. However, you have no experience with these treatments, and neither do your colleagues. On further enquiry, you discovered the following facts:

- Regarding **treatment C**, there are several very high-quality, independent studies comparing it to placebo. These have demonstrated significantly superior efficacy to placebo.
- Regarding **treatment D**, there are no studies measuring its efficacy.

All these treatments are very simple to use, and you are fully competent to do so. How confident are you in the efficacy of each treatment? Answer by moving the sliders below between “I’m certain it’s ineffective” and “I’m certain it’s effective”:

The participant is then asked to quantify their degree of confidence in the efficacy of each of the four treatments by clicking and moving a handle on a scale going from 0 – complete certainty that the treatment is inefficient, to 100 – complete certainty that the treatment is efficient. The four scales were placed right on top of each other, to allow direct visual comparison of their value, in an attempt to increase the reliability of the answers – see Figure 1. Indeed, to grasp the relative importance of scientific evidence and clinical experience for a given participant, we need to compare the values of the scales of each treatment (see section Statistical methods), so it is particularly important that these values are compared by the participant when answering. For instance, it is known that absolute judgement based on Likert scales are prone to response style bias (14), and we may expect the same bias with visual analog scale. Another method could have been to use a four-in-one multi-item scale, as described and studied in (15), but we chose to use this method instead for ergonomics and simplicity.

**Figure 1:**
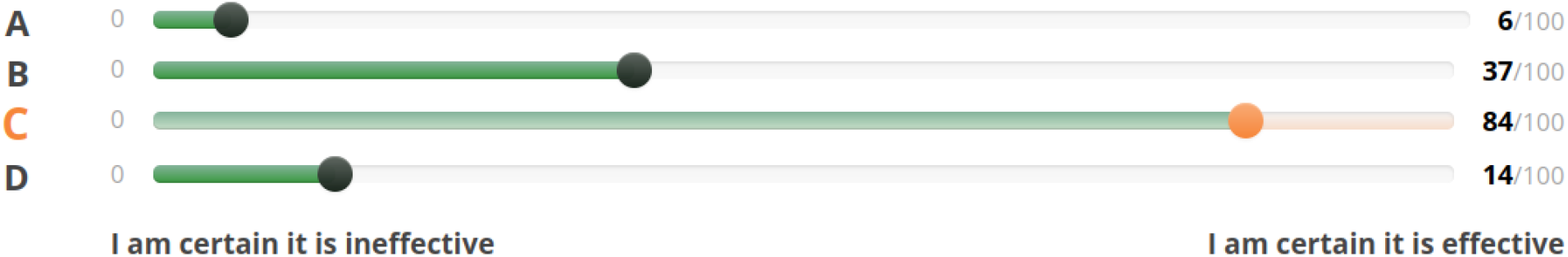
Sliders positioned by participants to answer the main question

After the main question, participants had the opportunity to comment on their answer, or to leave their email address for further discussion.

#### Demographic questions

In the second part of the survey, we asked the participants about their

- occupation: physiotherapist or physiotherapy student
- country of practice (for physiotherapists only)
- year of graduation (for physiotherapists only)
- year of study (for students only)
- previous participation in research paper production (published or unpublished)
- level of training in EBP: a score from 0 to 4, computed as the number of ‘Yes […]’ answers to these four questions:

QEBP1. Have you received teaching on the fundamental principles of EBP ? Answers: Yes, during initial training; Yes, during continuing training; Yes, during initial and continuing training; No; I don’t remember.

QEBP2. Have you received teaching on research on scientific databases (Pubmed and others) ? Same answers.

QEBP3. Have you received teaching on health research methodology (including different study designs: cross-sectional, longitudinal, randomized trial…) ? Same answers.

QEBP4. Have you received teaching on statistics ? Same answers.

The survey was created and administered using LimeSurvey Community Edition (LimeSurvey GmbH, n.d.) (16)

#### Ethics

This study is not considered as research involving human subjects according to the Jardé law in France. In respect of the European General Data Protection Regulation, all participants gave their consent to data processing by ticking an informative text box. Data was stored on a secured server of Grenoble Alpes University.

### Statistical methods

Let us first define two variables *Q+* and *Q*^−^, in equations (1) and (3) below.

Let *T*_*A*_,*T* _*B*_, *T*_*C*_, *T*_*D*_ ∈[0, 100] be the answer variables to the main question, relative to the treatments A,B,C, and D. For a given participant, we will quantify the importance of positive scientific evidence relative to clinical experience by defining the new variable *Q*^+^ by:

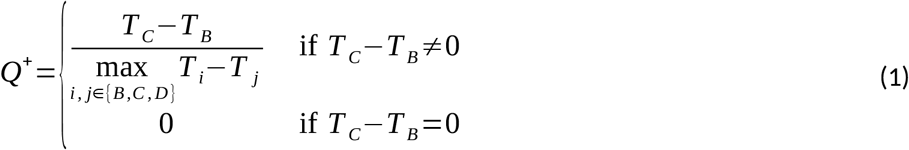

The reader may think that it would have been enough to define *Q*^+^= *T*_*C*_ −*T*_*B*_ to grasp the relative importance of positive scientific evidence and clinical experience. However, consider the two following possible answers to the main question, by two participants:

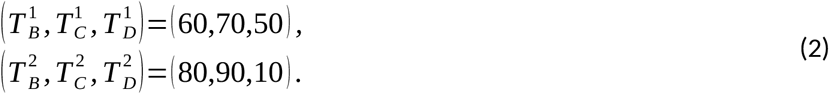

For these two participants, we have *T*_*C*_ −*T*_*B*_=10, which means that they both had more confidence in treatment C over treatment B, by a score of 10. Yet we argue that they don’t give the same importance to scientific evidence relative to clinical experience. For the first participant, we have 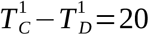. It means that positive scientific evidence made them increase their score for the reference treatment D by 20. Now, let us say we slightly modify the description of the treatment C in the main question, and make the positive scientific evidence a little bit less solid. For instance, instead of writing “there are several very high quality, independent studies […]”, we could write “there is one very high quality study […]”. Since the evidence is a little bit less solid, this participant may increase the score of D by only 75% of 20, so 15. They would then answer for this new scenario:

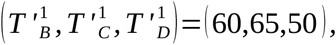

and so treatment C would still be the one which the first participant has the most confidence in. On the other hand, for the second participant we have 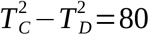. If we apply the same reasoning as before, the second participant would then answer for this new scenario:

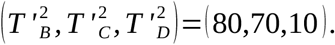

So treatment B would now be the one which the second participant has the most confidence in. We just showed that even though both participants initially preferred treatment C over treatment B by a score of 10, the robustness of this preference is different. The preference of the first participant for scientific evidence over clinical experience is stronger than that of the second participant. To measure the strength of the preference, we need some renormalization of *T*_*C*_ −*T* _*B*_, as proposed in equation (1). For these two participants, based on their answers from equation (2), we would have *Q*^+,1^=0.5 and *Q*^+,2^=0.125: we recover the fact that the first participant gives more importance to positive scientific evidence than the second participant, relative to clinical experience.

The interpretation of negative evidence will be analyzed in a coarser way with the categorical variable *Q*^−^,defined by:

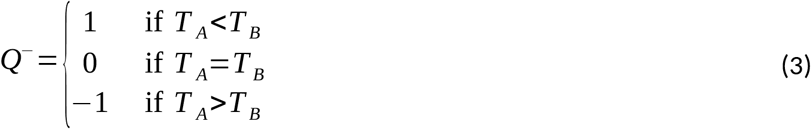

The results of the survey will be analyzed in a descriptive way through *Q*^+^ and *Q*^−^. To this end, we will classify the participants according to the value of *Q*^+^ as follows:

- If *Q*^+^ <−0.2, the participant has higher confidence in clinical experience than positive scientific evidence.
- If *Q*^+^ ∈[−0.2,0.2], the participant has equal confidence in clinical experience and positive scientific evidence.
- If *Q*^+^ >0.2, the participant has lower confidence in clinical experience than positive scientific evidence.

Let *EBP*∈{0,…, 4 } be the level of EBP training, determined by the number of ‘Yes […]’ answers to the questions regarding EBP training (QEBP1, QEBP2, QEBP3, and QEBP4). We will investigate the association between the outcome *Q*^+^ and the independent variable *EBP* with a bayesian ordered beta regression model, separately for students and physiotherapists. This regression model, introduced in (17), is an adequate model for modeling opinions expressed on a continuous bounded interval scale (and continuous data on a bounded interval in general). We also refer the reader to (18) for some less technical reader-friendly introduction to this model. We will compute the effect of changing the variable *EBP* from 0 to 4 on the expected value of *Q*^+^.

We will also investigate the association between the outcome *Q*^−^ and *EBP* with a multinomial logistic regression model.

Analysis was conducted using R (19) with Rstudio (20), and the packages ‘ordbetareg’ (17), ‘nnet’ (21), ‘marginaleffects’ (22).

### Sampling

We recruited a convenience sample of French-speaking physiotherapists practicing in France and Belgium, and French-speaking physiotherapy students. Participants were recruited through personal, professional and social network. Participant could answer the survey from December 15^th^, 2024, to February 14^th^, 2025.

It seems like a reasonable hypothesis to assume that the participants of our convenience sample hold a similar or higher opinion of scientific research than average, since they made the effort of answering this research survey. This means that, even though the distribution of the answers of the recruited participants might differ from the distribution of the answers of the target population, we may draw reasonable assumptions regarding the direction in which they should differ.

## Results

The questionnaire was completed by 263 students and 255 physiotherapists. The participants’ characteristics are summarized in Table 2.

**Table 2:**
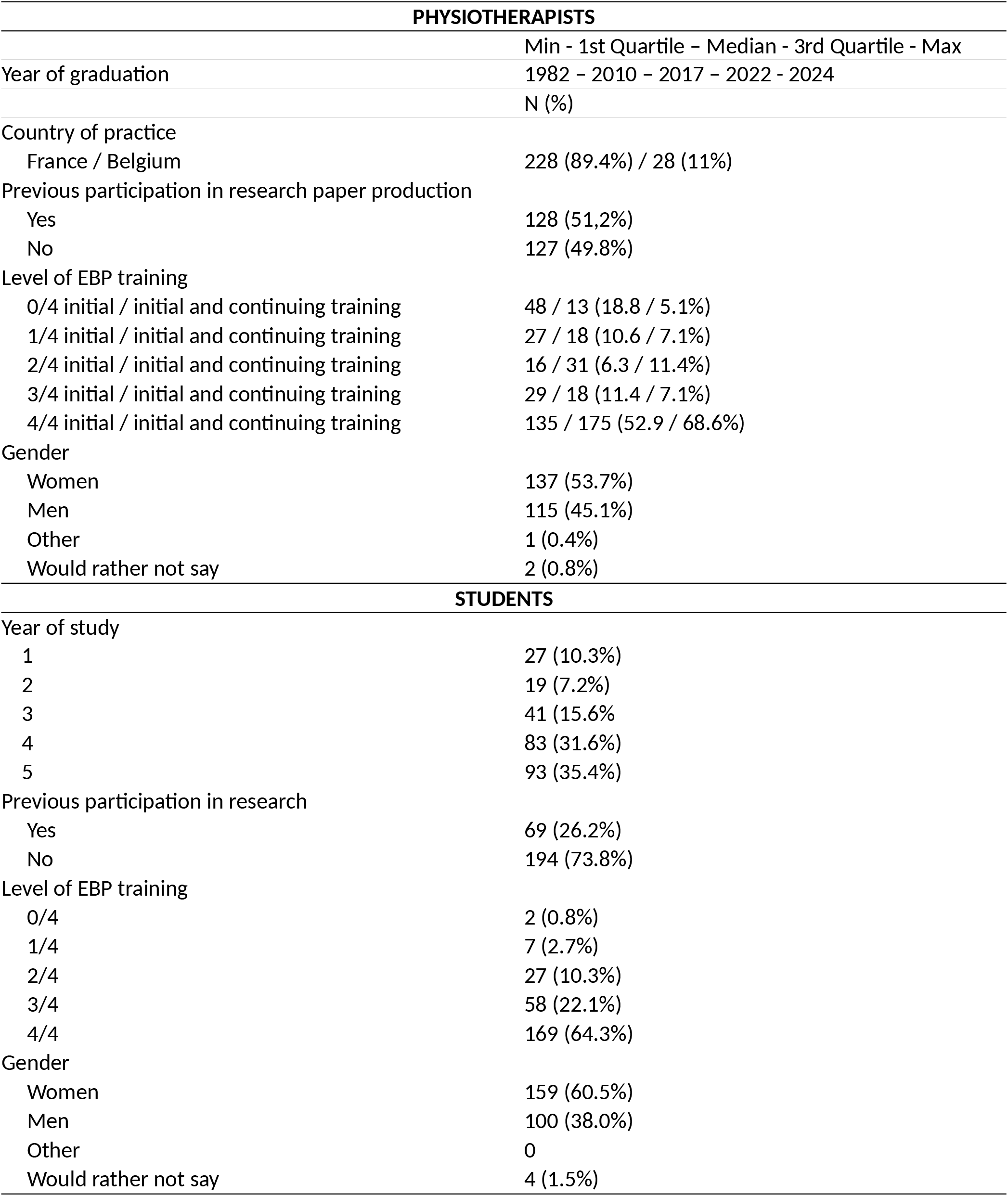
Participants’ characteristics.

The answers of five participants verified *T*_*C*_ <*T* _*D*_. It is highly surprising since it would mean that positive scientific evidence had a negative impact in their confidence in a treatment. Therefore, these answers were considered technical mistakes and were removed from the data. As it concerns only 1% of participants, doing so only marginally modifies the overall results towards higher confidence in scientific evidence.

The box plot of the scores T_A_, T_B_, T_C_ and T_D_ is given in Figure 2. Treatment C obtained the highest median score of 81, and treatment D obtained the lowest median score of 31. In between, treatment A obtained a slightly higher median score than treatment B, with 60 versus 50. However, in the following we will only be interested in comparing the treatments’ scores for each individual participant, via the outcomes *Q*^+^ and *Q*^−^.

**Figure 2:**
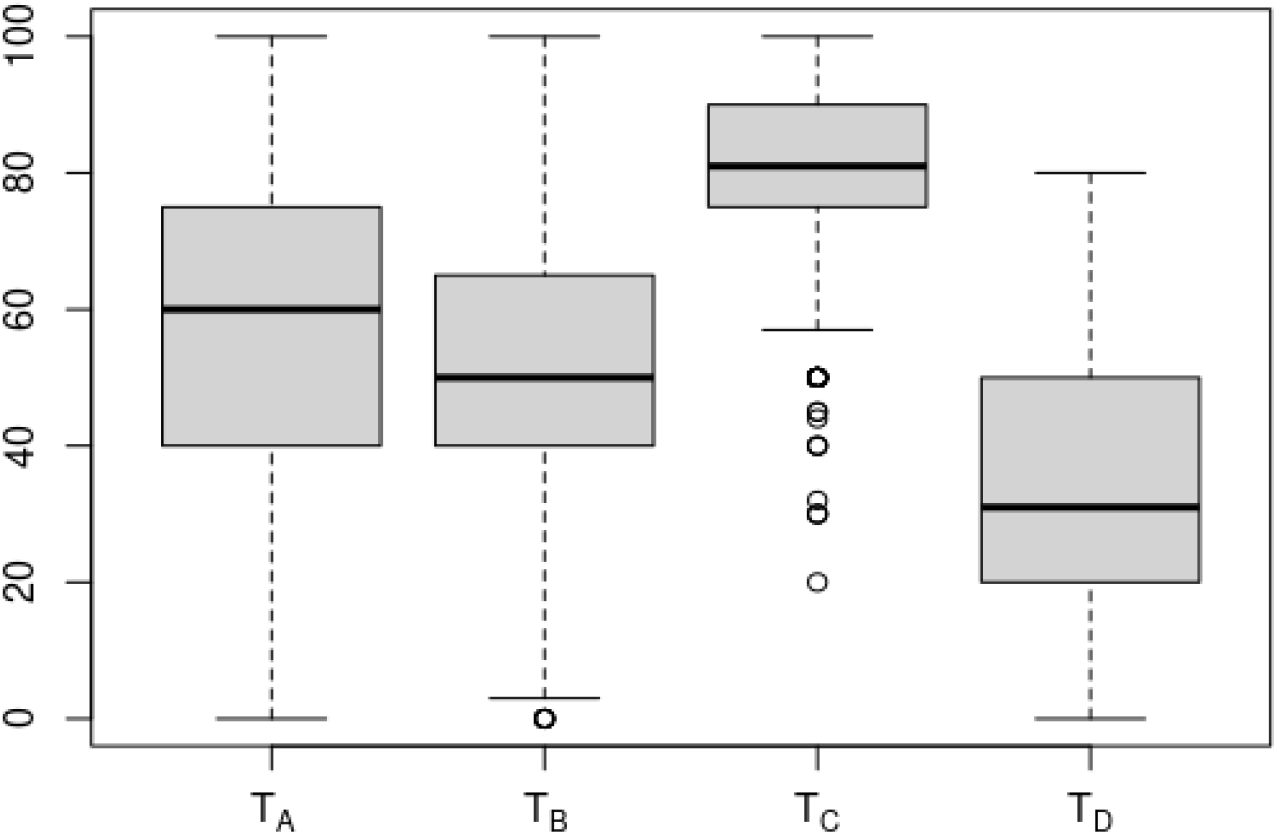
Box plot of the scores T_A_,T_B_,T_C_ and T_D_ (whiskers are 1.5 times the interquartile range)

The histogram of *Q*^+^ is displayed in Figure 3. The barplot of *Q*^−^ is displayed in Figure 4. The proportions of students and physiotherapists classified according to the values of *Q*^+^ and *Q*^−^ are given in Table 3, along with 95% confidence intervals.

**Table 3:** Proportions of students and physiotherapists classified according to the values of Q^+^ and Q^−^, with 95% confidence intervals (computed with the Wilson method for binomial proportions)

|  | Students |  | Physiotherapists |  |
| --- | --- | --- | --- | --- |
| $Q^+$ | | | | |
| $Q^+ < -0.2$<br>'more confidence in clinical experience than positive scientific evidence' | 7.2 % [4.7, 11.0] | 20.9 % [16.4, 26.2] | 5.5 % [3.3, 9.0] | 17.3 % [13.1, 22.4] |
| $Q^+ \in [-0.2,0.2]$<br>'equal confidence in clinical experience and positive scientific evidence' | 13.7 % [10.1, 18.4] | | 11.8 % [8.4, 16.3] | |
| $Q^+ > 0.2$<br>'lower confidence in clinical experience than positive scientific evidence' | 79.1 % [73.8, 83.6] | | 82.7 % [77.6, 86.9] | |
| $Q^-$ | Students | | Physiotherapists | |
| $Q^- = 1$<br>'negative evidence of treatment's efficacy decreased confidence' | 22.8 % [18.2, 28.3] | | 33.3 % [27.8, 39.3] | |
| $Q^- = 0$<br>'negative evidence of treatment's efficacy didn't impact confidence' | 20.2 % [15.7 , 25.4] | | 23.5 % [18.7, 29.1] | |
| $Q^- = -1$<br>'negative evidence of treatment's efficacy increased confidence' | 57.0 % [51.0 , 62.9] | | 43.1 % [37.2, 49.3] | |

**Figure 3:**
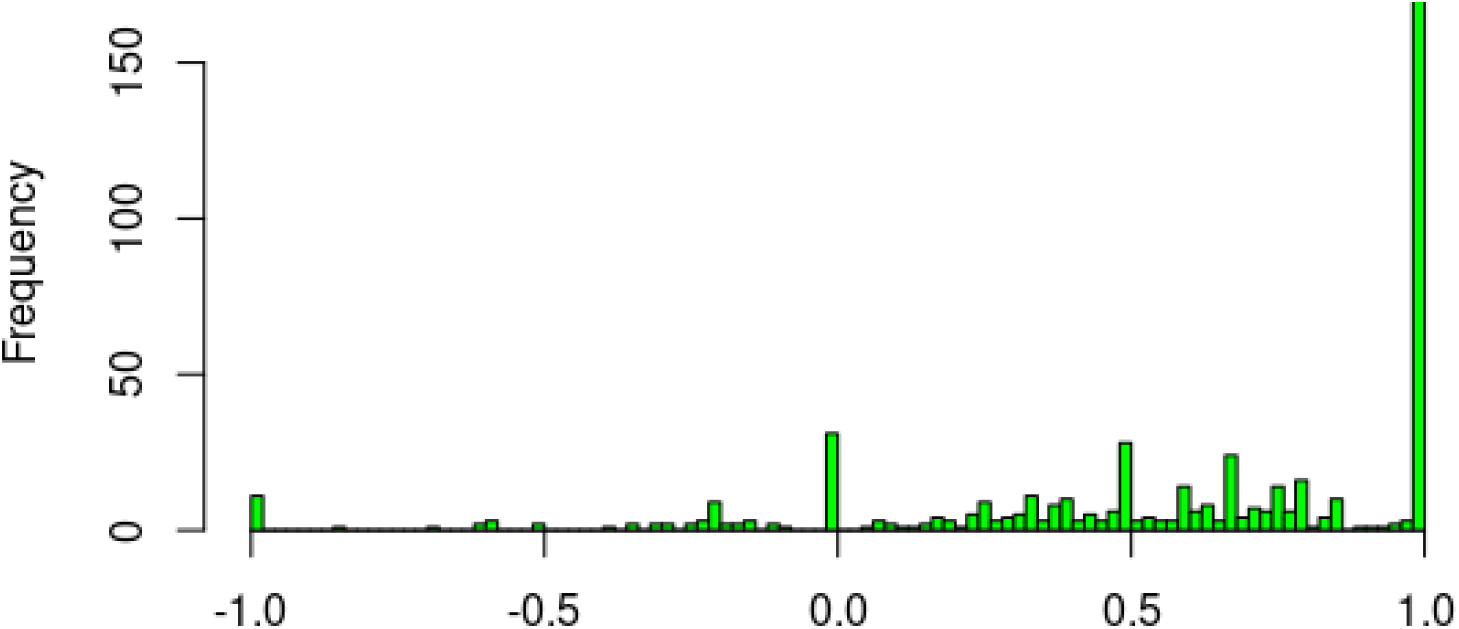
Histogram of Q^+^, for students and physiotherapists altogether

**Figure 4:**
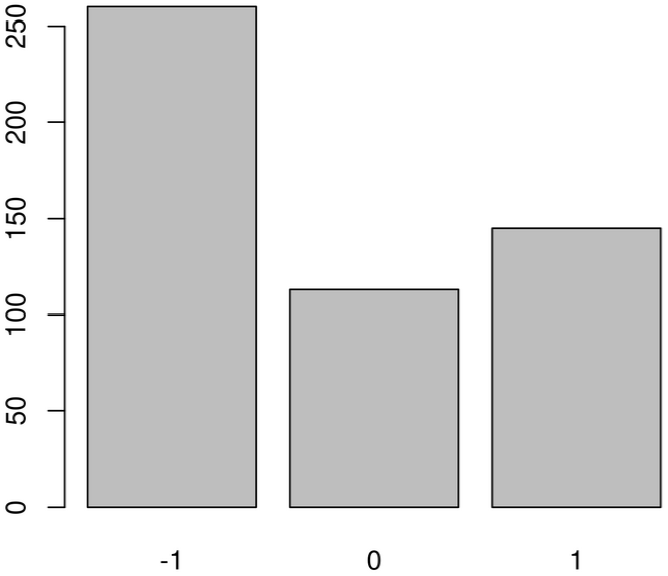
Barplot of Q^−^

### Positive evidence versus clinical experience

We classify most participants in the category ‘lower confidence in clinical experience than positive scientific evidence’ (*Q*^+^>0.2) (79.1% of students and 82.7% of physiotherapists). On the other hand, we classify 17% of physiotherapists and 21% of students in one of the two categories ‘equal confidence in clinical experience and positive scientific evidence’ (*Q*^+^ ∈[−0.2,0.2]), and ‘more confidence in clinical experience than positive scientific evidence’ (*Q*^+^ <−0.2).

### Negative evidence

For 22.8% of students and 33.3% of physiotherapists, knowing that a treatment’s efficacy had been studied by several very high quality studies and that this studies did not show any superiority to placebo, decreased their confidence in the treatment’s efficacy (*Q*^−^=1). For 20.2% of students and 23.5% of physiotherapists, it didn’t impact their confidence (*Q*^−^=0). For 57.0% of students and 43.1% of physiotherapists, it increased their confidence (*Q*^−^=−1).

### Association between Q^+^ / Q^−^ and EBP training level

The estimated parameters of each regression model are given in the supplementary material (SM2). Here we report the estimated effects of EBP training level, computed with the package ‘marginal effects’ in R (23).

In the ordinal beta regression model for students, the effect of increasing EBP training level from minimal to maximal (0 to 4) on the expected value of *Q*^+^ is –0.08, with a 95% confidence interval [–0.27, 0.14]. Given that *Q*^+^ takes its values in the interval [-1,1], effects of –0.27, –0.08, and 0.14 can arguably be considered as moderate negative, small negative, and small positive effects.

In the ordinal beta regression model for physiotherapists, the effect of increasing EBP training level from minimal to maximal (0 to 4) on the expected value of *Q*^+^ is 0.06, with a 95% confidence interval [-0.10, 0.24]. The effects –0.10, 0.06, and 0.24 can arguably be considered as small negative, small positive, and moderate positive effects.

In the multinomial logistic regression model, the a posteriori statistical power doesn’t allow us to conclude anything about the association between *Q*^−^ and EBP training level.

## Discussion

We used a novel method to investigate physiotherapists and physiotherapy students’ relative confidence in scientific evidence, positive and negative, and clinical experience, when evaluating a treatment’s efficacy. In a web-based survey, participants gave their degree of confidence in four fictitious treatments whose characteristics varied regarding scientific evidence and clinical experience.

The majority of the participants of our survey had higher confidence in positive scientific evidence than clinical experience. This is in line with previous studies evaluating physiotherapists attitudes towards evidence-based practice (8,13). Still, we classified 17% of physiotherapists and 21% of students in the categories ‘equal confidence in clinical experience and positive scientific evidence’ (*Q*^+^∈[−0.2,0.2]), and ‘higher confidence in clinical experience than positive scientific evidence’ (*Q*^+^ <−0.2) (Table 3). For these participants, a barrier to evidence-based practice is that they don’t agree with arguably the fundamental epistemological principle of evidence-based practice: (quality) scientific evidence is more reliable than clinical experience. In a systematic review on barriers to evidence-based practice in physiotherapy (8), ‘lack of interest’ was reported as a barrier by only 9% of participants. This shows that this proportion might have been underestimated by previous study designs.

Using an ordered beta regression model (17), we found that going from a minimal to a maximal EBP training level was associated with a variation of *Q*^+^ in the 95% confidence interval [-0.27, 0.14] for students, and [-0.10, 0.24] for physiotherapists. Given that *Q*^+^ ranges from –1 to 1, specific EBP training courses don’t seem to have any large effect on confidence in scientific evidence relative to clinical experience.

Regarding negative scientific evidence, our results are most intriguing. Let us remind the reader of the description of treatment A, regarding scientific evidence (see the Text Box above): ‘ there are several very high-quality, independent studies comparing it to placebo. These have not demonstrated superior efficacy to placebo’. As for treatment B, the description was ‘there are no studies measuring its efficacy’. Only 33% of physiotherapists and 23% of students had lower confidence in treatment A compared to treatment B (*Q*^−^=1). Moreover, 43% of physiotherapists and 57% of students actually had *higher* confidence (*Q*^−^=−1) (Table 3). Some of the latter participants had accepted to give their email address, so we had the opportunity to ask them for a brief justification of their answers. Moreover, some participants also commented on their answers in the comment box directly when answering the survey. All relevant comments are available in their original French versions on request. From this qualitative data, we inferred two ways of interpreting the information regarding treatment A, that led to increased confidence in this treatment:

- Interpretation 1: ‘ The description of treatment A implies that it is as effective as a placebo. Since placebos are known to be effective, treatment A is also effective.’
- Interpretation 2: ‘Treatment B doesn’t have any study on its efficacy. This warrants decreased confidence compared to treatment A.’

Interpretation 1 seems to be the most common one. It shows a strong misunderstanding of placebo effects and placebo controlled trials. Indeed, by definition of placebos, any treatment is at least as effective as its placebo equivalent – unless it has some specific harmful effects, but this wasn’t considered as a possibility in any justification given by the participants. Therefore, even though it is true that treatment A is as effective as ‘a placebo’ (its own placebo, actually), this is true of all treatments, and in particular of treatment B. So it shouldn’t justify increased confidence treatment A over treatment B.

Interpretation 2 implies that any study should increase one’s confidence in a treatment’s efficacy, regardless of its conclusion. Again, it could be argued that the description of treatment A implies that it was at least not found to be harmful, but none of the participants used that idea as a justification for that interpretation. It could also be argued that, since several studies were conducted on treatment A, but none on treatment B, the former must *a priori* be more effective than the latter – possibly because of a more plausible mechanism of action. This prior belief could then be so strong that it wouldn’t be completely modified by the studies’ contradictory conclusions. To justify this interpretation, one needs to give little credit to negative study findings.

For 24% of physiotherapists and 20% of students, negative scientific evidence didn’t increase nor decrease their confidence in a treatment’s efficacy (*Q*^−^ =0), (Table 3). One could hypothesize that these answers are due to the fact that the negative findings of studies were reported by the sentence ‘These have not demonstrated superior efficacy to placebo’ (SM1), which leaves a plausibility for some treatment’s efficacy that is meaningful but still not strong enough to be detected by these studies. However, none of the participants seem to have interpreted it this way, when justifying for their answers. On the contrary, our cautious description of negative findings was most of the time rephrased as a stronger statement such as ‘according to scientific literature, the treatment has no effect’, in the participants’ comments. Therefore, the most plausible interpretation is that evidence of inefficacy is simply ignored by these participants. This is in line with the results of (24), where the authors found, on a sample of physiotherapy students in India, that most participants would still recommend using a treatment after reading a Cochrane summary that indicates strong evidence of no benefit.

Results for physiotherapists and students were similar, but with an apparently slightly stronger tendency towards non-evidence based treatments for students. This difference could be explained by the highly likely fact that our students and physiotherapists samples are not representative of the underlying respective populations.

### Potential methodological improvement

In light of the results, we think the phrasing of the main question of the survey can be improved. For instance, a surprisingly high proportion (10%) of answers verified *T*_*B*_ <*T*_*D*_ (higher confidence in the treatment D than the treatment B). This is surprising as treatment D was designed to be a reference treatment on which very little is known, and treatment B was designed to be supported by clinical experience. However, because we wrote that treatment B had ‘long been used in your profession’, the absence of studies was interpreted more severely for treatment B than treatment D, which certainly appeared as a more recent treatment. This could have increased the proportion of answers with *Q*^+^=1. Therefore, removing the sentence ‘These have long been used in your profession.’ may improve the quality and interpretability of the results.

The results of the studies on the efficacy of treatment B (and C) were described by comparison to placebo (see the Text Box above). Interestingly, this led to the discovery of a strong misunderstanding of placebos and placebo controlled trials, and a non-expected interpretation of such negative trials. However, our primary intent was to investigate the interpretation of negative evidence in general. To this end, a phrasing that doesn’t refer to placebo could be preferred, even though it would make the statements less precise.

### Limitations

The answers may not reflect what participants actually think, because of the social desirability bias (25). (One participant did mention in a personal communication that it might have biased their answer.) As mentioned in the introduction, evidence-based practice is regarded as fundamental in physiotherapy, and this social norm might have influenced the answers.

The main question is cognitively demanding. Even though we encouraged participants to read and answer carefully, and tried to make the question as clear, simple and technically easy to answer as possible, the data might contain biased answers. For instance, we removed five answers because of the unlikely inequality *T*_*C*_ <*T* _*D*_. Moreover, one participant realized they had misread the question while emailing a justification for their answers.

Our convenience sampling method doesn’t allow us to get a representative sample of the target population. However, participants might value scientific research more than non-participants, since they made the effort to answer the survey. This hypothesis is also consistent with the fact that 50% of the physiotherapists participants had previously participated in the production of a scientific research paper (Table 2). It allows us to generalize some of our results outside of our sample.

### Future research

Specific EBP training courses seem to have no to at most moderate effect on agreement with the fundamental principle of evidence-based practice that high-quality research is more robust than clinical experience. It would be useful to identify some knowledge or skill associated with such agreement. For instance, the knowledge of the most common reasons why a clinician may be misled to conclude that a treatment they used was effective (such as placebo effect, regression to the mean, wording effect, …), could theoretically decrease one’s confidence in clinical experience to evaluate a treatment’s efficacy.

## Conclusions

High confidence in clinical experience may have been an underestimated barrier to evidence-based practice in previous studies, as 17.3% [13.1, 22.4] of physiotherapist and 20.9% [16.4, 26.2] of student participants had equal or lower confidence in positive scientific evidence than clinical experience. General EBP training seems to have no to at most moderate effect on this relative confidence. The interpretation of negative findings of placebo-controlled trials show that most physiotherapists and physiotherapy students misunderstand placebos or ignore negative study findings. This could be an important barrier to evidence-based practice.

## Data Availability

Data presented in this study is available in an anonymized version in the supplementary material file SM3_anonymized_data.csv

