## Supplementary materials SM1 and SM2 for "How do scientific evidence and clinical experience influence physiotherapists’ confidence in a treatment’s efficacy ? A survey of French-speaking physiotherapists"

**SM1 : English version of the questionnaire**

***Part 1/2 : Main question***

**This question forms the core of the questionnaire. Please take the time to read and answer it carefully. Thank you very much!**

You are treating a new patient, Mr. X. To treat his health problem, you are considering 4 possible treatments (A, B, C and D), which have the following characteristics.

- **Treatments A and B**: These have long been used in your profession. You are accustomed to using one or the other, and you observe an improvement in the condition of the patients with whom you use these treatments. What's more, your colleagues also use them and are satisfied. However, one of them recently pointed out to you that there are no studies to prove their effectiveness. On further enquiry, you discovered the following facts:
  - Regarding **treatment A**, there are several very high-quality, independent studies comparing it to placebo. These have not demonstrated superior efficacy to placebo.
  - Regarding **treatment B**, there are no studies measuring its efficacy.
- **Treatments C and D:** A colleague recently told you about these two options, which, according to some sources, are showing interesting results. However, you have no experience with these treatments, and neither do your colleagues. On further enquiry, you discovered the following facts:
  - Regarding **treatment C**, there are several very high-quality, independent studies comparing it to placebo. These have demonstrated significantly superior efficacy to placebo.
  - Regarding **treatment D**, there are no studies measuring its efficacy.

All these treatments are very simple to use, and you are fully competent to do so. How confident are you in the efficacy of each treatment? Answer by moving the sliders below between “I'm sure it's ineffective” and “I'm sure it's effective”:

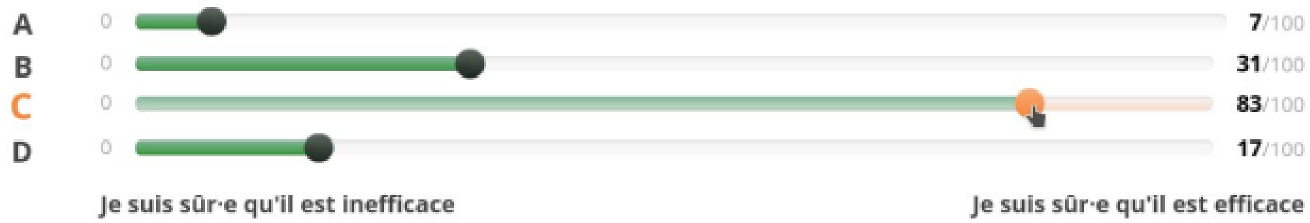


***Part 2/2 : sociodemographic questions***

• You are a:

- *physiotherapist*

- *physiotherapy student*

• In which country(ies) do you work presently? [For physiotherapists only]

- *France*

- *Belgium*

- *Other*

• What year did you obtain your physiotherapist degree ?  [For physiotherapists only]

• Which institution did you graduate from ?  [For physiotherapists only]

• What is your current year of study ? [For students only]

• What institution are you studying at ? [For students only]

• Have you received teaching on the fundamental principles of EBP (or « evidence-based practice ») ?

- *Yes, during initial training [For physiotherapists only]*

- *Yes, during continuing training [For physiotherapists only]*

- *Yes, during initial and continuing training [For physiotherapists only]*

- *Yes [For students only]*

- *No*

- *I don't remember*

• Have you received teaching on research on scientific databases (Pubmed and others) ?

- *Yes, during initial training [For physiotherapists only]*

- *Yes, during continuing training [For physiotherapists only]*

- *Yes, during initial and continuing training [For physiotherapists only]*

- *Yes [For students only]*

- *No*

- *I don't remember*

• Have you received teaching in health research methodology (including different study designs : cross-sectional, longitudinal, randomized trial…) ?

- *Yes, during initial training [For physiotherapists only]*

- *Yes, during continuing training [For physiotherapists only]*

- *Yes, during initial and continuing training [For physiotherapists only]*

- *Yes [For students only]*

- *No*

- *I don't remember*

•  Have you received teaching in statistics ?

- *Yes, during initial training [For physiotherapists only]*

- *Yes, during continuing training [For physiotherapists only]*

- *Yes, during initial and continuing training [For physiotherapists only]*

- *Yes [For students only]*

- *No*

- *I don't remember*

• Have you participated in the production of a scientific research article (published or unpublished)?

- *Yes*

- *No*

• What gender do you identify with ?

- *Woman*

- *Man*

- *Other*

- *I would rather not say*

*SM2 : Regression models*

***Ordered beta regression model***

In this model (1), the distribution of $Q^{+}$ is a function of the variable $EBP$ and some regression parameters, defined by :

$$L_{Q^{+}} = \begin{cases} 1 - g(\alpha + \beta \cdot EBP - k_0)\,\delta_0 \\ + \ \left(g(\alpha + \beta \cdot EBP - k_0) - g(\alpha + \beta \cdot EBP - k_1)\right) Beta\left(g(\alpha + \beta \cdot EBP), \phi\right) \\ + \ g(\alpha + \beta \cdot EBP - k_1)\,\delta_1 \end{cases} \quad (1)$$

where $g$ is the inverse logit function, $\delta_0$ and $\delta_1$ are Dirac masses at 0 and 1, $\alpha,\ \beta,\ k_1,\ k_2$, and $\phi$, are parameters of the model to be estimated from the data, and Beta is the usual Beta distribution.

The following results were obtained using the default priors of the ordbetareg() function of the 'ordbetareg' package in R. For all variables, Rhat = 1.00.

**Physiotherapists :**

Regression Coefficients:
$\alpha$ : 0.75, 95%CI=(0.40,1.09)
$\beta$ : 0.03, 95%CI=(-0.06,0.13)

Further Distributional Parameters:
$\phi$ : 5.88, 95%CI=(4.70,7.17)
$k_0$ : -2.82, 95%CI=(-3.68,-2.10)
$k_1$ : 1.40, 95%CI=(1.22,1.60)

**Students :**

Regression Coefficients:
$\alpha$ : 1.00, 95%CI=(0.55,1.43)
$\beta$ : -0.05, 95%CI=(-0.17,0.07)

Further Distributional Parameters:
$\phi$ : 5.09, 95%CI=(0.49,4.18)
$k_0$ : -3.02, 95%CI=(0.42,-3.90)
$k_1$ : 1.59, 95%CI=(0.09,1.43)

***Multinomial logistic regression models***

Physiotherapists :

| $Q^{-}$ | term | estimate | std.error | statistic | p.value | conf.low | conf.high |
|---|---|---|---|---|---|---|---|

| | | | | | | | |
|---|---|---|---|---|---|---|---|
| -1 | (Intercept) | 0.582 | 0.487 | 1.19 | 0.233 | -0.374 | 1.54 |
| -1 | EBP | -0.0944 | 0.135 | -0.698 | 0.485 | -0.360 | 0.171 |
| 0 | (Intercept) | 1.05 | 0.479 | 2.19 | 0.0283 | 0.111 | 1.99 |
| 0 | EBP | -0.441 | 0.140 | -3.16 | 0.00160 | -0.715 | -0.167 |

Students:

| Q⁻ | term | estimate | std.error | statistic | p.value | conf.low | conf.high |
|---|---|---|---|---|---|---|---|
| -1 | (Intercept) | 2.47 | 0.776 | 3.18 | 0.00147 | 0.947 | 3.99 |
| -1 | EBP | -0.442 | 0.212 | -2.08 | 0.0372 | -0.858 | -0.0262 |
| 0 | (Intercept) | 0.228 | 0.985 | 0.232 | 0.817 | -1.70 | 2.16 |
| 0 | EBP | -0.0976 | 0.268 | -0.365 | 0.715 | -0.622 | 0.427 |
